# How young people experienced Long COVID services: a qualitative analysis

**DOI:** 10.1101/2024.09.12.24312643

**Authors:** Olivia Taylor, Georgia Treneman-Evans, Madeleine Riley, Joanne Bond-Kendall, Katharine Pike

## Abstract

**Objective:** We aimed to evaluate how acceptable paediatric Long COVID services were to patients and clinicians. This paper focuses on how acceptable Long COVID services were to paediatric patients.

**Design:** This study was explorational. Semi-structured qualitative interviews with 13 paediatric patients were used to understand the experiences of patients with Long COVID.

**Setting:** Participants were recruited from specialist paediatric services in the Southwest of England from June 2022 to September 2023.

**Patients:** Participants were children and young people (CYP) aged 11-17 years old with a Long COVID diagnosis who accessed the specialist services in the Southwest of England.

**Results:** Four themes were reported. Accessing specialist clinics helped CYP to feel validated, they appreciated consulting with clinicians who were knowledgeable about Long COVID and empathetic. CYP found comfort in knowing other CYP were experiencing Long COVID. CYP wanted to be proactive in their Long COVID management, appreciating regular appointments and the opportunity to learn about their condition. CYP desired normality, and therefore sought flexible appointment times, online appointments, and reasonable adjustments. CYP found the waiting times to access Long COVID services were too long.

**Conclusions:** Our results stress the importance to CYP of several features of the care received in the specialist clinics. These relate to the experiences of CYP with Long COVID but potentially extend to CYP with other conditions, particularly long-term and/or poorly understood conditions. The results support creating community-based support groups for CYP with long-term medical conditions, providing online flexible appointments, offering early reasonable adjustments for school and providing quicker access to specialist clinics.

**Key messages/Summary box:** *What is already known on this topic:* The existing literature which explores adult experiences of Long COVID services is in agreement with this study in multiple ways. Firstly, it appears that clinician validation is very important to the clinician patient relationship (1-5). Peer support and building a Long COVID community for patients is also valued by individuals with Long COVID (1, 3, 5, 6), as are online appointments (1,3). Finally, long waiting lists to access Long COVID clinics are a barrier to care (1, 3). This can be mitigated to some extent by providing sign-posting or self-management resources while waiting for clinic access (3). However, the literature regarding paediatric experiences of Long COVID is limited. Two papers discuss the impact of a lack of reasonable adjustments on school (7,8) and the importance of validation through peer support (7).

*What this study adds:* To the best of our knowledge, there are no studies on Long COVID services for paediatric patients that highlight how patients experienced management of their condition. Our study highlights the patient’s desire to be proactive in their management, including being given opportunities to learn about Long COVID.

*How this study might affect research, practice or policy:* This study highlights recommendations for clinical practice. It is recommended to consider the following when designing specialist paediatric services for CYP with Long COVID: CYP with complex medical conditions want clinicians to validate their experience by acknowledging that their symptoms are real and their concerns are valid. Community-based groups for CYP experiencing the same condition are valued by patients, as are online flexible appointments, liaison with schools to provide early reasonable adjustments and rapid access to services for CYP. Future research should explore how these recommendations positively impact patients.

## Introduction

‘Ongoing symptomatic COVID-19’ is defined as signs and symptoms of COVID-19 lasting four to twelve weeks post COVID-19 infection. ‘Post-COVID-19 syndrome’ is defined as signs and symptoms which occur during infection with COVID-19 or after, which then continue for more than twelve weeks and cannot be explained by another diagnosis (9). Both ‘Ongoing symptomatic COVID-19’ and ‘Post-COVID-19 syndrome’ are often referred to as Long COVID (9). Estimates from meta-analyses suggest the prevalence of Long COVID symptoms after infection with COVID-19 in children and young people (CYP) was 23-% (10, 11).

After the COVID-19 pandemic, CYP with Long COVID symptoms were managed by Long COVID hubs. In the South of the UK, CYP experiencing fatigue as a symptom of Long COVID were also managed by a specialist fatigue service. In the South of the UK, the Long COVID hub closed in 2023, but patients who were under the care of a specialist service received continued care. General Practitioner’s (GP’s) can now refer CYP with suspected Long COVID directly to a specialist fatigue service or other speciality. The medical consequences of COVID-19 on CYP are well documented (insert ref) but young people’s experiences of Long COVID managed in specialist services has not been well examined. This paper evaluates the experiences of CYP managed by the South West Long COVID services. This data can be more widely extrapolated to help improve service user experience for CYP who engage in other specialist clinics.

## Methods

### Design

A mixed-method approach was employed. REDCap (a secure online database) was used to collect quantitative data from at least 50 adolescents aged 11-17-years-old. Adolescents were asked to answer questions relating to their wellbeing and quality of life. In addition, qualitative data was collected from at least 12 adolescents aged 11-17-years-old who were invited to a semi-structured one-to-one interview with the first or third author. These interviews explored the experiences of receiving a Long COVID diagnosis and of management within the specialist Long COVID clinics. This paper reports the qualitative data only.

### Recruitment

Patients from two specialist paedatric services were invited to the study, those from the South West Long COVID hub and the specialist fatigue service. Staff within these services identified and approached eligible patients at their clinic appointments and asked their consent to be contacted by a member of the study team. Patients with a diagnosis of Long COVID who were between 11 and 17 years old were eligible. Exclusion criteria for the study included children who were unable to complete questionnaires or take part in interviews, for example those who due to extreme fatigue or pain were unable to be interviewed for 15 minutes. Patients who consented to contact were then contacted via email by the study team to arrange an online or in-person interview.

### Participants

Interviews have been assigned a letter A-M in order of appearance in the text, with the letter having no link to the interview. We aimed to use purposive sampling to capture a wide range of ages and genders in the study. However, recruitment was lower than expected so purposive sampling was not feasible. Instead, we invited participants to an interview if they provided consent to be contacted about an interview. Thirteen participants completed an interview between September 2022 and September 2023. Interviews were carried out by the first author and the second author. Based on participant preference, 12 of the 13 interviews were conducted over Zoom and one interview at one of the specialist services in a non-clinical setting. The interviews were no longer than 45 minutes. Participants were given the option of whether they would like their parent/carer present during the interview. Most interviews were audio recorded using Zoom software. The interviews followed a structured topic guide which was piloted with PPI (patient and public involvement) members and adapted as suggested for the study. At the beginning of the interviews, participants were asked about their initial symptoms of COVID-19 infection and their ongoing symptoms (i.e., their Long COVID). This was to gain an understanding of the participants’ experiences. Participants were then asked further questions exploring their experiences of attending the Long COVID services. No repeat interviews were carried out. Field notes were made after the interviews had been carried out for analytic purposes. Transcripts were not returned to participants for comment or correction. 1 participant declined participating in the study, and 1 participant noted they were too ill to participate in the study.

### Qualitative analysis

Thematic analysis was used (Braun and Clarke, 2006). The first author manually transcribed each interview after it had taken place, therefore familiarising herself with the data. The first author then read each interview at least twice to immerse herself in the data. The first author generated initial codes from the data using NVIVO software; these initial codes were a way of organising the data by assigning transcript quotations to categories. Once 13 interviews had been completed, the first author felt saturation had been reached so interviews did not continue. The second author coded 7 of the 13 interviews to ensure accurate and reliable coding was achieved. The research team then met via Microsoft Teams to discuss any discrepancies with the coding. Following this meeting the first author developed themes from the data.

### Ethics approval

Ethical approval was obtained in May 2022 from Health and Care Research Wales (HCRW).

## Results

### Participants

Thirteen young people completed a one-to-one semi-structured interview. Of the 13 participants, 8 provided baseline data regarding their age, gender and ethnicity. The ages of these 8 participants, ranged between 12 and 16 years old. Of the 8 participants who reported baseline data, 7 identified as female and 1 as male. Of the 8 participants who reported baseline data, all identified as White (Table S1).

During the initial COVID-19 infection, the most common symptoms reported were fatigue, headaches and brain fog (Tables S2 & S3).

Consistent with previous studies in CYP, fatigue was the most common reported symptom of Long COVID (12). Also common were headaches, dizziness/fainting, and musculoskeletal symptoms (Tables S4 & S5).

### Generation of codes

The first author developed initial codes including symptoms of initial COVID-19 infections, symptoms of Long COVID, what is good about the Long COVID service, what could be improved about the Long COVID service, what is the effect of Long COVID on school, would you recommend the Long COVID service to other children with Long COVID. When the second author co-coded 7 of the 13 interviews, no discrepancies in codes generated were found.

### Generation of themes

The first author developed themes from the coded data. There were four themes 1) validation of experience, 2) young people wanted to be proactive in their Long COVID management, 3)young people desired normality 4) waiting for versus getting help (Table S6). Participants were not asked for feedback on the findings.

### Theme 1 Validation of experience

#### Subtheme 1 Young people appreciated healthcare professionals who had knowledge of Long COVID

Early in the COVID-19 pandemic, knowledge of long-term consequences of COVID-19 infection was limited. CYP reported mixed reactions from healthcare professionals. Some CYP reported feeling dismissed by their general practitioner (GP), with one young person describing how her GP thought her symptoms of Long COVID were all in her head. Another young person noted that healthcare professionals at the Long COVID clinic were more understanding of Long COVID than other professionals. In general, participants reported a more positive experience in the Long COVID clinics. The participants appreciated that the healthcare professionals at the clinics were knowledgeable about Long COVID, and empathised with the young people’s symptoms. The participants valued seeing healthcare professionals who validated their lived experiences of having Long COVID, who were empathetic, kind, and most of all believed them. When asked about their experience of using the Long COVID clinics, one participant answered:

> *‘Participant: Yeah it’s been fine easy to talk to um and very understanding of the situation which is the difference of speaking to someone whose specialised in it over the past couple of years to just a general doctor or GP.’ (Interview A: did not provide demographic information)*

Another participant described her experience of using the Long COVID clinics as validating:

> *‘Participant: I think it was just because the doctors had been so kind of dismissive it’s all in your head. There’s not a lot. Everyone goes through it. And I was suffering with pain every day and I still am. And it’s like for a while I found that really hard and like even when we moved, I found it really hard just to be able to go to the doctors because I was just like, I don’t want them to be telling me it’s all in my head again. So, when we had the doctors or the CFS people turn around and go, oh my God, you’ve been through so much…It made me cry at the end of it because I was very much like, I felt, listened to’ (Interview B: Female, White)*

#### Subtheme 2 Young people were validated in the knowledge that other CYP were using the Long COVID clinic

Knowing other young people were using the Long COVID clinic also validated the CYP’s lived experiences, reducing the feelings of isolation associated with having a new and little understood condition. One participant stated they would find a support group for young people with Long COVID helpful. They expressed that it would be useful to share experiences of Long COVID with other CYP. The young people liked the sense of community the clinics provided. When asked about their experience of the Long COVID clinics, one participant expressed relief at knowing other CYP were going through the same experience:

> *‘Participant: And then, just to hear that I’m not the only one going through it, like, there are other people just don’t worry. It was just like it was like a weight off the shoulder.’ (Interview F: did not provide demographic information)*

When asked what would improve their experience of using the Long COVID clinics, one participant suggested setting up a Long COVID support group for CYP.

> *‘Participant: um I think, um, obviously hospitals should have areas, because we don’t really have a Long COVID area in the hospital do we?*… *Which I think that would be a better idea to be able to go there and actually face to face other people who have it and all go together and tell each other things like help and what not.’ (Interview C: did not provide demographic data)*

### Theme 2 Young people wanted to be proactive in their Long COVID management

Participants wanted their symptoms of Long COVID to improve; they therefore valued the feeling of being proactive in managing their Long COVID symptoms. This was achieved through having regular clinic appointments, and learning about Long COVID in order to find new ways of managing their symptoms.

CYP expressed this sentiment as:

> *‘Participant: it’s quite relieving to know that people are you know doing something.’ (Interview E: Male, White)*
>
> *‘Participant: I felt like there was going to be at least someone that’s going to kind of look into making a change.’ (Interview B: Female, White)*

#### Subtheme 1) Regular clinic appointments helped young people manage their symptoms of Long COVID

Having regular clinic appoints helped the young people feel proactive in managing their Long COVID symptoms. Young people found regular clinic appointments reduced anxiety or uncertainty around managing symptoms; If they were having difficulties with their management plans, they could target these issues sooner and therefore get better faster. Participants described how they found regular clinic appointments helpful for their recovery:

> *‘Participant: sometimes I can still feel very exhausted, but it’s definitely helped and it has calmed me down a lot because sometimes I get very stressed wondering how long this is going to go on for… but it’s the fact that every two weeks I’m up here it calms me down a lot.’ (Interview D: Female, White)*
>
> *‘Participant: …because we’ve got a shorter period of meetings now, so instead of doing it every two weeks, it’s every like every one week instead… It’s a lot more helpful and when I’m struggling with something that’s going on in my mind and it’s feels like it’s a kind of circle… you kind of like get trapped and sometimes it’s nice to yeah, have that support; when like if it was the week that I wasn’t talking to her, it would be quite hard to her and go, oh, this happened last week, but being able to have it and go, this is happening right now. It helps me kind of move forward a bit more.’ (Interview B: Female, White)*

#### Subtheme 2) Learning about Long COVID helped young people manage their symptoms of Long COVID

Another way young people found they could be proactive in managing their symptoms was to learn about Long COVID and use this knowledge to aid their recovery. Participants appreciated the extra steps the staff went to in order to make sure they understood the content of the clinic appointments. Participants described how they appreciated receiving information about Long COVID and their management plans:

> *‘Participant: umm I liked how they kind of given me lots of information because it’s nice knowing that you know all these things can be ruled out…it’s nice having lots of people with different skillsets so it’s nice you know having a wide range of information.’ (Interview E: Male White)*
>
> *‘Participant: Yeah. So, a lot of the time they kind of send like emails and debriefs after it because I obviously suffer from brain fog…but like they’ve kind of helped kind of digest it after and give the leaflets to kind of go through PowerPoints that explains certain bits for like my age or what we could make better so yeah, that’s definitely been a big help.’ (Interview B: Female, White)*

### Theme 3 Young people desired normality

#### Subtheme 1) Young people valued flexible appointments to help have a sense of normality

Most participants expressed how putting school and social life first was important to them. They valued appointment times to suit their schedule. Clinic appointments took place at times suggested by the participants, and this enabled CYP to ensure their appointments didn’t clash with school timetabling. When asked if the appointment times worked with their schedule, participants responded:

> *‘Participant: yes yeah they did and we planned it and they’ve always kept it in the afternoon which is really helpful for me.’ (Interview G: Female, White)*
>
> *‘Participant: Yeah it was all catered around me, so whenever I had free periods I would organise one. Basically we just planned the date and time that suited us both and it always worked. I didn’t miss any school for it.’ (Interview H: Female, White)*

#### Subtheme 2) Young people valued online appointments to help with their Long COVID management

Participants favoured appointments held online since this reduced the time and cost of commuting to appointments. Participants also appreciated that online appointments were less impactful on their energy levels than travelling to physical appointments. When participants were asked if they would rather have in person appointments, they responded:

> *‘Participant: …I don’t think I would have gained any, I think it would have been worse because I would have had to travel for it and then it would have been a bigger thing and probably more energy consuming.’ (Interview H: Female, White)*
>
> *‘Participant: The online appointments are really helpful. If I’d had to travel to [clinic location] because I’m up in the [home location], yeah, that would have triggered my fatigue before I even got to the appointment. So don’t stop the online appointments because it just makes it so much easier if you’re on one end of the country, you don’t have to drive all the way to the other end to get advice.’ (Interview I: Female, White)*

#### Subtheme 3) The clinics helped the young people gain reasonable adjustments for school

The participants also appreciated the positive impact the Long COVID clinics had on their school education because healthcare professionals could provide a letter with information about the participant’s Long COVID to their school. One participant also attended Cognitive Behavioural Therapy (CBT) which provided management strategies to help them sit their school exams.

Participants noted:

> *‘Participant: From the appointments my mum gets some letters sometimes and she forwards them to the school which means they have all the information and it means if there’s any like really big issues I know who to talk to yeah.’ (Interview E: Male, White)*
>
> *‘Participant: …The CBT was really helpful with getting through my exams. I wouldn’t have been able to do those without the management strategies that that gave me…But we definitely needed the clinic. We wouldn’t have been able to get here without the support they gave with school.’ (Interview I: Female, White)*

### Theme 4: Waiting for versus getting help

#### Subtheme 1) The wait to access Long COVID clinics was too long

Nearly all of the participants felt that the waiting list to be seen at the Long COVID clinics was too long. The participants described how they had to wait to be referred into the Long COVID service from their GP or local hospital, the participants then discussed how they had to wait after being referred for their first clinic appointment.

The time it took for participants to access the clinics ranged from a few months to over a year. Participants reported that they had little support or advice about managing their Long COVID for long periods of time, while experiencing long COVID symptoms. Many participants were not able to attend school full time while waiting to access Long COVID clinics, and did not have medical exemption or support to miss school, access reduced timetabling, extenuating circumstances, or support with examinations. When participants were asked how they would improve their experience of the Long COVID clinics, they answered:

> *‘Participant: If they could like figure out how to make it go a bit faster, because I had to wait years before even getting referred.’ (Interview J: Female, White)*
>
> *‘Participant: Because it’s always really busy, so it’s like six months to get an appointment with the paediatrician, and then we only see him once every six months, and every time we have to convince him it’s actually Long COVID again. And then it’s another 3-6 months to get referred to the fatigue clinic and then that’s another couple of months to get onto the CBT. So, it doesn’t feel like there’s enough people doing it for the amount of people who need it, which is no fault of the doctors.’ (Interview I: Female, White)*

#### Subtheme 2) Young people wanted information on how to help manage their symptoms whilst waiting for an appointment

When asked what improvements could be made to the specialist clinics, participants said it would be helpful to be seen by clinics more quickly. Participants put forward some suggestions which could help young people wanting interim support whilst on a waiting list. Participants suggested giving CYP on the waiting list guidance about what they can do to help manage their Long COVID symptoms, and also giving more guidance to support CYP between clinic appointments. One participant suggested an NHS inform page on paediatric Long COVID would be helpful. Whilst there is an NHS inform page on Long COVID management for fatigue, low mood and depression and joint pain, this is not tailored specifically to meet the needs of the paediatric Long COVID population (13,14,15) :

> *‘Participant: UM. Well, before we sort of got round to the fatigue clinic. For treatment with fatigue, there wasn’t sort of at least that I know of any like NHS page with recommendations. There might be, and I just haven’t found it, but we didn’t really know what to do with fatigue management. We didn’t know what to do with CBT. We didn’t know how to manage my dizziness or headaches or any of my symptoms, really.’*
>
> *‘Participant: Yeah. And also, just general tips and advice, so don’t overdo it. Don’t get into a boom-and-bust cycle. And when you’re doing too much and then you crash and repeat over and over again. Like ways to help yourself rest like meditation or CBT or anything. As far as I’m aware, I might be incorrect in this, but there is no place that you can go where all that information is sort of in one place on the NHS website.’ (Interview I:Female, White)*

#### Subtheme 3) Young people wanted the clinics to communicate with their school whilst they were waiting

Having interim support for Long COVID whilst awaiting clinic management was especially important for school; participants noted that without communications from the clinic to the school, it could be hard to gain extra support in school :

> *‘Participant: Or if there was someone you could contact to get those things, because if you like, got Long COVID couple of months before your exam, the chances that you’re going to get seen, see if specialist in that time before your exam to be able to take that exam with the mitigating circumstances are very slim, so either you don’t sit your exam, or you sit it and really really struggle.’ (Interview I: Female, White)*

## Discussion

### Summary of results

This study reports the experiences of 13 CYP who received specialist support for Long COVID. Participant CYP appreciated healthcare professionals at the Long COVID clinics validating their experiences. They also felt validated in their experiences of Long COVID by knowing that other CYP were affected by Long COVID too. Participants reported wanting to be proactive in manging their symptoms of Long COVID, so appreciated regular clinic appointments and being educated about Long COVID. CYP wanted to prioritise their school education and normal lives, so appreciated clinic appointments being scheduled to suit their timetable, and clinic appointments being online; CYP appreciated the Long COVID clinics helping school to conduct reasonable adjustments, such as help with timetabling and examinations. Overall, the Long COVID clinics were well received, but most CYP had concerns about length of wait to access Long COVID clinics.

### Recommendations: clinical

The experiences and perspectives of young people who received support from the Long COVID clinics could be used to inform the design of similar services, for example in supporting CYP with other long-term, complex medical conditions. Participants reported feeling validated during consultations because the health professionals used language that expressed empathy and compassion; healthcare professionals should consider the impact of language and behaviours upon CYP’s perceptions that their symptoms are real and their concerns valid.

CYP found knowing other CYP were also experiencing symptoms of Long COVID validated their experiences; services could consider creating community-led support for patients such as online support groups.

These findings also highlight the benefit of helping patients feel proactive in managing symptoms of complex long-term conditions. Healthcare professionals could facilitate young people to feel proactive by providing them with regular appointments, offering information about their illness, and offering research updates potentially in the form of a patient newsletter.

Healthcare professionals supporting young people with complex long-term conditions, should consider CYPs the impact of long-term conditions upon education, friendships and activities and aim to minimise further negative impacts in these areas, supporting CYP where possible to engage with the activities that are important to them. One way to facilitate this is through organising flexible online clinic appointments.

As suggested by our findings, young people with complex long-term conditions need easy and early access to specialist support. Their symptoms need to be addressed in a timely manner in order to minimise the impact on their education, working lives, physical and mental health. Where services cannot be expanded to meet this need, a mitigation might be providing information and guidance for young people awaiting their specialist clinic appointment by signposting online and other freely available resources. The importance of clinics who manage long term complex conditions for young people being able to explain to schools that a young person is seeking medical help, and may need adaptations to their school timetable, and examinations before they are formally assessed should not be underestimated. A multi-agency approach between specialist clinics and schools may be necessary to provide children and young people with holistic support in all areas of their lives.

### Recommendations: research

The findings of this paper highlight the importance of flexible data collection, for example conducting online interviews at a time that works well for participants or meeting them face to face at a convenient location and offering breaks during an interview. Further research could explore how the clinical recommendations made in this paper might impact patients’ experiences and outcomes.

### Strengths of the study

This study was able to reach data saturation and conduct a rigorous thematic analysis to better understand the experiences of 13 CYP with Long COVID in the South of England. Our participants had a heterogenous experience of COVID-19 infection, ranging from mildly symptomatic to hospitalised, this captured a wider range of experiences. However, the views showed strong agreement and similarity in experiences, and therefore we have been able to make recommendations for future clinical practice.

### Limitations of the study

Participants had different initial experiences of COVID 19 infection ranging from mildly symptomatic to hospitalised. This may have influenced how much they needed the support of the clinics and their evaluation of them. The interviews we conducted did not all take place at the same time point in a participant’s journey of Long COVID, for example, some participants had developed Long COVID symptoms recently whereas others had developed Long COVID symptoms a long time ago. This might have influenced participants response to questions, for example when asking if they would recommend the clinics to other young people with Long COVID, participants seemed more likely to say yes if they had been using the clinic services for a longer time period. Another limitation of our study is the lack of diversity within our cohort, most of the participants gender and ethnicity identified as female and reported their ethnicity as White.

## Supporting information

COREQ checklist

## Data Availability

All data produced in the present work are contained in the manuscript or supplementary materials.

