## Supplementary Materials for "How young people experienced Long COVID services: a qualitative analysis"

### Baseline demographics of participants

Table 1: Baseline demographics of participants

| Interview | Gender given | Ethnicity given |
| --- | --- | --- |
| F – did not provide baseline data |  |  |
| A – did not provide baseline data |  |  |
| E | Male | White |
| G | Female | White |
| C – did not provide baseline data |  |  |
| L | Female | White |
| D | Female | White |
| J | Female | White |
| K – did not provide baseline data |  |  |
| B | Female | White |
| M – did not provide baseline data |  |  |
| G | Female | White |
| I | Female | White |

### Symptoms of acute COVID-19 infection

Table 2: Symptoms which participants reported during their initial COVID-19 infection

Symptom reported by participants

Fatigue

Headaches

Brain Fog

Fever

Sensory disturbance

Nausea or vomiting

Dizziness or fainting

Cough

Limb weakness or pain

Purple feet

Sore throat

Rash

Fast heart beat

Loss of appetite

Table 3: Symptoms of acute COVID-19 infection supporting evidence

| Symptom reported by participants | Supporting evidence |
| --- | --- |
| Fatigue | - ‘Participant: Umm, so when I first got it on Christmas day I was just very tired, had like a really bad headache, because all I wanted to do was just sleep, but because the headache was so bad I couldn’t really get off to sleep. And then that kind of like stayed for maybe the next couple of weeks it’s always kind of there when I get tired.’ - ‘Participant: umm feeling tired just wanted to be in bed basically and just very run down.’ - ‘Participant: um fatigue, brain fog, um mainly those two I mean there were others but that was it mainly.’ - Participant: yeah a cough, a headache, or I just felt a bit tired.’ - ‘Interviewer: So we’ve started recording our discussion so lets start by talking about when you had coronavirus tell me a bit about that…’   ‘Participant: Uh I was um just like really really tired like constantly and I would like only get up just to eat and then just go back to sleep again.’   - ‘Participant: Yeah, sluggish, not being able to smell, taste; tasting horrible.’ - ‘Participant: Ohh OK, that makes sense now. Quite tiring. Couldn't concentrate that much, really lots of headaches. Felt sick. And that was it really, I don’t think there was much more was there? - ‘Participant: Umm, I think to be honest I don’t really remember. I’ve had it three times so far and it’s kind of been a different experience each time. I think it was, it kind of just took me out, it was quite draining, I think I had, I had a cough I think, yeah I was in bed for I think like a week or something.’ - ‘Participant: I first got COVID in I think it was October 2020. It wasn't like your standard COVID infection. I didn't lose my taste or smell, but I had a really bad headache, and I couldn't get out of bed for about a week. My taste didn't go away, but it changed, so food started tasting different. The only thing I really wanted to eat at that point was tomato soup.’   ‘Participant: At the start it was headaches and fatigue and then the dizziness as well and difficulty thinking.’ |
| Headaches | - ‘Participant: Umm, so when I first got it on Christmas day I was just very tired, had like a really bad headache, because all I wanted to do was just sleep, but because the headache was so bad I couldn’t really get off to sleep. And then that kind of like stayed for maybe the next couple of weeks it’s always kind of there when I get tired and it was just part of the COVID thing just that headache. - ‘Participant: Well when I first had COVID it wasn’t that bad I wasn’t very ill I think the most I had was just a headache’ - ‘Participant: I had a bad headache as well too’ - ‘Participant: Ohh OK, that makes sense now. Quite tiring. Couldn't concentrate that much, really lots of headaches. Felt sick. And that was it really, I don’t think there was much more was there?’ - ‘Participant: I first got COVID in I think it was October 2020. It wasn't like your standard COVID infection. I didn't lose my taste or smell, but I had a really bad headache, and I couldn't get out of bed for about a week. My taste didn't go away, but it changed, so food started tasting different. The only thing I really wanted to eat at that point was tomato soup.’   ‘Participant: At the start it was headaches and fatigue and then the dizziness as well and difficulty thinking.’ |
| Brain Fog | - ‘Participant: Umm I had a heart rate of 180 Interviewer: oh wow Participant: and the paramedics thought that I was going to go into arrythmia or cardiac arrest’ ‘Interviewer: yeah’ ‘Participant: so they took me straight in uhh and I stayed for 12 days and my symptoms were I couldn’t stand up, I couldn’t sit up, I nearly fainted a couple times in the hospital, I was sick, I was really dehydrated I don’t know if that’s a symptom but I was put on IV again, um, I had brain fog again, I had the hospital team and I barely did anything oh yeah I had purple feet as well which I still have I think it’s now referred to as COVID toes.’ - ‘Participant: um fatigue, brain fog, um mainly those two I mean there were others but that was it mainly.’ - ‘Participant: Ohh OK, that makes sense now. Quite tiring. Couldn't concentrate that much, really lots of headaches. Felt sick. And that was it really, I don’t think there was much more was there?’ - ‘Participant: At the start it was headaches and fatigue and then the dizziness as well and difficulty thinking.’ |
| Fever | - ‘Participant: not getting your temperature quite right, too hot or too cold quite often.’ - ‘Participant: um I couldn’t stand the first there were two different sort of symptoms so August 2020 I had a fever of 40 degrees for five days, I was sick, I fainted, I had a rash, um, uhh, trying to think what else there was now, yeah and I was ill like that for 5 or 6 days then I went to the hospital and I stayed in for 4 nights and then the second time I went in I was rushed in by ambulance.’ |
| Sensory disturbance | - ‘Participant: um I couldn’t taste anything, I couldn’t smell anything uh I had a really sore and gross throat as well.’ - ‘Participant: Yeah, sluggish, not being able to smell, taste; tasting horrible.’   ‘Participant: It tasted like sick.’ ‘Participant: Yeah, like rotten eggs and everything smelled horrible. I couldn't stand like in a room with anything like smelling, cooking, candles, couldn't stand with any sprays because it just smelled so bad.’   - ‘Participant: I first got COVID in I think it was October 2020. It wasn't like your standard COVID infection. I didn't lose my taste or smell, but I had a really bad headache, and I couldn't get out of bed for about a week. My taste didn't go away, but it changed, so food started tasting different. The only thing I really wanted to eat at that point was tomato soup.’ |
| Nausea or vomiting | - ‘Participant: umm I had a heart rate of 180’ ‘Interviewer: oh wow’ ‘Participant: and the paramedics thought that I was going to go into arrythmia or cardiac arrest’ ‘Interviewer: yeah’ ‘Participant: so they took me straight in uhh and I stayed for 12 days and my symptoms were I couldn’t stand up, I couldn’t sit up, I nearly fainted a couple times in the hospital, I was sick, I was really dehydrated I don’t know if that’s a symptom but I was put on IV again, um, I had brain fog again, I had the hospital team and I barely did anything oh yeah I had purple feet as well which I still have I think it’s now referred to as COVID toes.’   ‘Participant: um I couldn’t stand the first there were two different sort of symptoms so August 2020 I had a fever of 40 degrees for five days, I was sick, I fainted, I had a rash, um, uhh, trying to think what else there was now, yeah and I was ill like that for 5 or 6 days then I went to the hospital and I stayed in for 4 nights and then the second time I went in I was rushed in by ambulance.’   - ‘Participant: I felt really sick aswell like all the time.’ - ‘Participant: Ohh OK, that makes sense now. Quite tiring. Couldn't concentrate that much, really lots of headaches. Felt sick. And that was it really, I don’t think there was much more was there?’ |
| Dizziness or fainting | - ‘Participant: fainting whenever I had water on my head like a bath or a shower I would faint after it I couldn’t wash myself or anything.’ - ‘Participant: umm I had a heart rate of 180’ ‘Interviewer: oh wow’ ‘Participant: and the paramedics thought that I was going to go into arrythmia or cardiac arrest’ ‘Interviewer: yeah’ ‘Participant: so they took me straight in uhh and I stayed for 12 days and my symptoms were I couldn’t stand up, I couldn’t sit up, I nearly fainted a couple times in the hospital, I was sick, I was really dehdryated I don’t know if that’s a symptom but I was put on IV again, um, I had brain fog again, I had the hospital team and I barely did anything oh yeah I had purple feet as well which I still have I think it’s now referred to as COVID toes.’   ‘Participant: um I couldn’t stand the first there were two different sort of symptoms so August 2020 I had a fever of 40 degrees for five days, I was sick, I fainted, I had a rash, um, uhh, trying to think what else there was now, yeah and I was ill like that for 5 or 6 days then I went to the hospital and I stayed in for 4 nights and then the second time I went in I was rushed in by ambulance.’   - ‘Participant: At the start it was headaches and fatigue and then the dizziness as well and difficulty thinking.’ |
| Cough | - ‘Participant: um I always struggle with this one because it was so long ago now but it was like umm so there was a dry cough.’ - ‘Participant: yeah a cough, a headache, or I just felt a bit tired.’ - ‘Participant: Umm, I think to be honest I don’t really remember. I’ve had it three times so far and it’s kind of been a different experience each time. I think it was, it kind of just took me out, it was quite draining, I think I had, I had a cough I think, yeah I was in bed for I think like a week or something.’ |
| Limb weakness or pain | - ‘Participant: and then obviously the major one was loosing the legs.’ |
| Purple feet | - ‘Participant: umm I had a heart rate of 180’ ‘Interviewer: oh wow’ ‘Participant: and the paramedics thought that I was going to go into arrythmia or cardiac arrest’ ‘Interviewer: yeah’ ‘Participant: so they took me straight in uhh and I stayed for 12 days and my symptoms were I couldn’t stand up, I couldn’t sit up, I nearly fainted a couple times in the hospital, I was sick, I was really dehydrated I don’t know if that’s a symptom but I was put on IV again, um, I had brain fog again, I had the hospital team and I barely did anything oh yeah I had purple feet as well which I still have I think it’s now referred to as COVID toes.’ |
| Sore throat | - ‘Participant: um I couldn’t taste anything, I couldn’t smell anything uh I had a really sore and gross throat as well.’ |
| Rash | - ‘Participant: um I couldn’t stand the first there were two different sort of symptoms so August 2020 I had a fever of 40 degrees for five days, I was sick, I fainted, I had a rash, um, uhh, trying to think what else there was now, yeah and I was ill like that for 5 or 6 days then I went to the hospital and I stayed in for 4 nights and then the second time I went in I was rushed in by ambulance.’ |
| Fast heart beat | - ‘Participant: umm I had a heart rate of 180’ ‘Interviewer: oh wow’ ‘Participant: and the paramedics thought that I was going to go into arrythmia or cardiac arrest’ ‘Interviewer: yeah’ ‘Participant: so they took me straight in uhh and I stayed for 12 days and my symptoms were I couldn’t stand up, I couldn’t sit up, I nearly fainted a couple times in the hospital, I was sick, I was really dehydrated I don’t know if that’s a symptom but I was put on IV again, um, I had brain fog again, I had the hospital team and I barely did anything oh yeah I had purple feet as well which I still have I think it’s now referred to as COVID toes.’ |
| Loss of appetite | - ‘Participant: I lost my appetite for a while didn’t I.’ |

### Symptoms reported of Long COVID

Table 4: Symptoms reported of Long COVID

Symptom reported by participants

Fatigue

Headaches

Dizziness or fainting

Limb pain or weakness

Difficulties with sleep

Abdominal pain or vomiting or nausea or reflux

Sensory issues

Brain fog

Appetite loss

Changes to menstruation

Mental health

Blackouts

Vision problems

Vertigo

Fever

Table 5: Symptoms of Long COVID

| Symptom reported by participants | Supporting evidence |
| --- | --- |
| Fatigue | - ‘Participant: no, but when people will be like so why can’t you come out and play football like it’s not going to be that hard it’s like no because Long COVID you get tired it’s not anything personal but I’ll get too tired and then I’ll faint on you and do you really want me to faint in front of you.’ - ‘Participant: After that like the long COVID signs only really kicked in for me especially about a month or two after umm because we hadn’t really noticed once we’d gone into lockdown and stuff we weren’t really doing much so we just thought it would be tiredness me being sort of routine shifted being a teenager anyway being quite lethargic but then realised when I started having to do stuff it was just so much more hard work than it was for anyone else that’s when we sort of realised it.’   ‘Participant: Yeah so the main one was the fatigue that was the one that took over.’   - ‘Participant: Yeah um I think I think this is fatigue they said um laten um by bedtime is normally meant to be around 9pm laten it to around you know 10 ish 11 and then wake up earlier at 8am will mean I have better quality sleep I think that was something that they said.’   Participant: I feel a little bit tired afterwards but I think I mean it’s worth feeling that tired finding out the information.’  ‘Participant: so I have to explain it and be like I get tired a lot of the time’  ‘Participant: I often say well I’ve got Long COVID which means I get tired a lot of the time and I struggle with lots of exercise and stuff doing lots of activities’   - ‘Participant: I’ve still got fatigue I still haven’t gone to school in two and a half years’   ‘Participant: no I was continuously unwell, the first hospital admission we didn’t know what was wrong so I was still going up and down the stairs and I was pretty much crashing daily I felt horrendous and then obviously after the second time we moved my bed downstairs where we’re still sleeping um we I couldn’t stand up I couldn’t get up off the sofa I needed help being escorted to the toilet which is like we have a downstairs toilet it’s like 5 steps away from the sofa.’  ‘Participant: so even my OT was saying she was struggling to keep up with all of my symptoms because a lot of them aren’t anything to do with chronic fatigue they’re purely to do with Long COVID so like when I had the gastro problems um I couldn’t eat anything or I ate but it was incredibly painful I was very weak I couldn’t lift my head off the sofa so there was no way I was going to be like practising climbing the stairs and going out for walks and building up my energy envelope when I couldn’t even stand up?’   - ‘Participant: yeah like I’m on a part time timetable in school because of my um sleep because my energy is really bad I can’t manage it’ - ‘Participant: uh yeah, I wasn’t getting out as much as I used to because I was either too tired or just not feeling up to it and it affected my eating habits and I just kind of stopped doing a lot of the things that I used to do.’ - ‘Participant: then it’s like some days I feel really ill I can barely get out of bed, like this morning’ ‘Interviewer: what were you feeling this morning?’ ‘Participant: oh I was just feeling really exhausted like normally I can get up to go downstairs and eat my food but this morning it was like I couldn’t get out of bed and then when I did I was just getting really bad pains in my legs and my head would hurt a lot yeah.’   Participant: yeah every since I’ve just got either a small headache or a migraine yeah really bad.’   - ‘Participant: Well the first time I was unwell for months and months and I couldn’t do a lot of school I was just so tired.‘   ‘Interviewer: Is there anything else you would like to share with me about your experience of having Long COVID or chronic fatigue syndrome?’ ‘Participant: Um well what happen is I basically just keep on sleeping and even if I’ve got loads of sleep when I wake up I’m still tired and if I don’t decide to push myself out of bed I’ll just keep on sleeping and it’s really annoying.’  ‘Interviewer: It sounds like you would like to have more energy, you would like to feel less tired.’ ‘Participant: Yeah because a lot of the time I have to cancel things with my friends because I feel so tired and it’s a bit annoying.’  ‘Participant: And that's I think that's the hardest thing at the moment is just learning that even the most smallest tasks can make you so exhausted.’  ‘Had fatigue, the chronic fatigue that I've now got. So, I was really exhausted doing things like walking up and down the stairs was really tough for me and still actually quite is.’   - ‘Participant: Then the bad week. If it's really bad, three. If it’s so so like feeling sick, quite tired, probably 5.’ - ‘Participant: and yeah it was just like fatigue and uh kind of full body tiredness and mental tiredness not like because I get tired before but it wasn’t the same it was full exhaustion.’ - ‘Participant: And so after about two weeks, I went back to school. I knew something wasn't right because I was feeling exhausted and headachy all the time. But it was sort of manageable. I'd go to school. I'd come home at normal time. And when I got home, I would go straight to bed because I was so tired I couldn't stay up. I was struggling to keep up with homework cause I was going to bed straight after school, so I didn't have time to do it. And then that lasted about half a term. And then at Christmas, something changed and everything sort of got a lot worse all at once. So the headaches got a bit worse and I stopped sleeping almost completely. So at nights where I was getting absolutely no sleep, there was nights where I was getting about two hours sleep. That was about the standard I was getting at that point. Uh, good night sleep was 4 hours then.   ‘Participant: At the start it was headaches and fatigue and then the dizziness as well and difficulty thinking. Those all persisted, the sense of taste came like a day couple of days. UM. And then the dizziness got worse so there was a point where if I sat down and stood up, I'd have to wobble for a couple of moments before I'd be stable, there was a point where I stood up and I was like, OK, for a couple of seconds and then I just completely collapsed. But that might have been a medication that I was on at that point. The dizziness got better first. The headaches are still present. The fatigue is still present. The insomnia is still present. The brain fog is still present, but less, it's either not being triggered as much because I'm doing subjects that it likes more or it has, well, it has got a bit better too.’ |
| Headaches | - ‘Participant: uhh headaches, dizziness, nausea, umm sort of funny stomach gut sort of area that was never amazing umm but yeah the fatigue and the tiredness was the main one.’ - ‘Participant: But I want to say around the 4th one is when the long covid really started to happen which was um not good um it was there was a lot of different symptoms other than just the normal ones from covid’ ’ ‘Participant: stomach aches uh burning feet the reflux thing where my chest it was burning and it felt like it was going up and down my chest headaches um a lot of stuff like that.’ - ‘Participant: Then it’s like some days I feel really ill I can barely get out of bed, like this morning’ ‘Interviewer: what were you feeling this morning?’ ‘Participant: Oh I was just feeling really exhausted like normally I can get up to go downstairs and eat my food but this morning it was like I couldn’t get out of bed and then when I did I was just getting really bad pains in my legs and my head would hurt a lot yeah.’   Participant: yeah every since I’ve just got either a small headache or a migraine yeah really bad.’   - ‘Participant: And so after about two weeks, I went back to school. I knew something wasn't right because I was feeling exhausted and headachy all the time. But it was sort of manageable. I'd go to school. I'd come home at normal time. And when I got home, I would go straight to bed because I was so tired I couldn't stay up. I was struggling to keep up with homework cause I was going to bed straight after school, so I didn't have time to do it. And then that lasted about half a term. And then at Christmas, something changed and everything sort of got a lot worse all at once. So the headaches got a bit worse and I stopped sleeping almost completely. So at nights where I was getting absolutely no sleep, there was nights where I was getting about two hours sleep. That was about the standard I was getting at that point. Uh, good night sleep was 4 hours then.’   ‘Participant: At the start it was headaches and fatigue and then the dizziness as well and difficulty thinking. Those all persisted, the sense of taste came like a day couple of days. UM. And then the dizziness got worse so there was a point where if I sat down and stood up, I'd have to wobble for a couple of moments before I'd be stable, there was a point where I stood up and I was like, OK, for a couple of seconds and then I just completely collapsed. But that might have been a medication that I was on at that point. The dizziness got better first. The headaches are still present. The fatigue is still present. The insomnia is still present. The brain fog is still present, but less, it's either not being triggered as much because I'm doing subjects that it likes more or it has, well, it has got a bit better too.’ |
| Dizziness or fainting | - ‘Participant: and the one still with the long covid is like if I get tired or I’ve done too much I tend to faint after it ummm’   ‘Participant: no, but when people will be like so why can’t you come out and play football like it’s not going to be that hard it’s like no because Long COVID you get tired it’s not anything personal but I’ll get too tired and then I’ll faint on you and do you really want me to faint in front of you.’   - ‘Participant: Uhh headaches, dizziness, nausea, umm sort of funny stomach gut sort of area that was never amazing umm but yeah the fatigue and the tiredness was the main one.’ - ‘Participant: Um I still feel dizzy I still feel faint that was getting better but obviously with the recent infection quotation marks that has come back and now I’m nearly unable to stand without being dizzy.’ - ‘Participant: no it’s just like sometimes if I get my blurred vision I can feel a bit dizzy or it’s like sometimes in class if it’s too loud or I feel a bit stressed it can kind of cause a bit dizzy.’ - ‘Participant: At the start it was headaches and fatigue and then the dizziness as well and difficulty thinking. Those all persisted, the sense of taste came like a day couple of days. UM. And then the dizziness got worse so there was a point where if I sat down and stood up, I'd have to wobble for a couple of moments before I'd be stable, there was a point where I stood up and I was like, OK, for a couple of seconds and then I just completely collapsed. But that might have been a medication that I was on at that point. The dizziness got better first. The headaches are still present. The fatigue is still present. The insomnia is still present. The brain fog is still present, but less, it's either not being triggered as much because I'm doing subjects that it likes more or it has, well, it has got a bit better too.’ |
| Limb pain or weakness | - ‘Interviewer: Yeah definitely it sounds like you had a lot of different symptoms at various times and tell me a bit more about what happened with your legs ,if you don’t mind’ ‘Participant: um yeah absolutely, yeah they went really tingly and I just thought maybe it’s me not doing any exercise because I was on the sofa all the time, and then I completley lost it and when people was touching them I just couldn’t feel it umm and then it kept going and going and I think it’s for like 2 months straight I had no feeling in them whatsoever.’ - ‘Participant: But there were a lot of other symptoms below that so there was umm muscle aches get sort of muscle aches all over your body especially in your hands, and your fingers, umm so that was quite bad one.’ - ‘Participant: I’ve got musculoskeletal pain’ - ‘Participant: But I want to say around the 4th one is when the long covid really started to happen which was um not good um it was there was a lot of different symptoms other than just the normal ones from covid’ ‘Participant: stomach aches uh burning feet the reflux thing where my chest it was burning and it felt like it was going up and down my chest headaches um a lot of stuff like that.’ - ‘Participant: Then it’s like some days I feel really ill I can barely get out of bed, like this morning’ ‘Interviewer: What were you feeling this morning?’ ‘Participant: oh I was just feeling really exhausted like normally I can get up to go downstairs and eat my food but this morning it was like I couldn’t get out of bed and then when I did I was just getting really bad pains in my legs and my head would hurt a lot yeah.’   ‘Participant: Yeah I also get aching pains in my legs and arms a lot of the time like most of the time like this morning and yesterday I woke up with bad aching pains and it just stays.’ |
| Difficulties with sleep | - ‘Participant: But that’s another thing with the Long COVID actually that I’ve had so like sleeping can actually be difficult no matter how tired you are it’s just more like it’s your body that’s tired rather than your brain so you’re wanting to sort of stay awake but your bodies telling you no’ ‘Interviewer: yeah that must be really difficult um so how long did you struggle with the sleep issues for?’ ‘Participant: still do to be honest which um is probably like I can put some of it on the long covid part of it but then I have to also look at myself and think I’ve just had my summer it’s probably my sleep patterns a bit out of place anyway.’ - ‘Interviewer: yeah is sleep um like a problem for you then?’ ‘Participant: yeah yeah it’s a very big problem’ ‘Interviewer: in what way?’ ‘Participant: um waking up in the middle of the night with um headaches and stuff is very very common’ ‘Interviewer: um’ ‘Participant: usually takes a while to get to sleep because waking up in the night takes a while to get back to sleep aswell and then I’ll either not go back to sleep or sleep like a deep sleep until like 11 o clock’ ‘Interviewer: right yeah so a bit all over the place’ ‘Participant: yeah’ ‘Interviewer: yeah, and how long has that been going on for for you?’ ‘Participant: a very long time’   ‘Interviewer: So just going back to the sleep thing what would you say your waking times are like at the moment when are you going to sleep and waking up?’ ‘Participant: so usually like 2 in the morning um and then if I manage to get back to sleep then it’s yeah it’s usually completley random from like so 2 in the morning is when I wake up not to like wake up and do stuff obviously just waking up in the morning I’ll get back to sleep hopefully about an hour to get to sleep um and then mum will try and wake me up around or she’ll give me a wake up call at around sort of 9 and then half an hour later I’ll probably still be in bed uh she’ll call me again um and that happens until about 11 and then 11 is when I have to try and really get up for the day.’ ‘Interviewer: right okay so that’s kind of started when the covid all started the sleep disturbances yeah and do you find that you’re quite tired in the day with that do you need to take naps and things like that?’ ’Participant: yeah like I’m on a part time timetable in school because of my um sleep because my energy is really bad I can’t manage it.’   - ‘Participant: um, my sleep is a lot worse than it used to be, um and it’s not just from Long COVID because I also have migraines which affect my sleep pattern as well. But um my sleep has always been kind of bad but its gotten worse since I got Long COVID.’ - ‘Participant: And so after about two weeks, I went back to school. I knew something wasn't right because I was feeling exhausted and headachy all the time. But it was sort of manageable. I'd go to school. I'd come home at normal time. And when I got home, I would go straight to bed because I was so tired I couldn't stay up. I was struggling to keep up with homework cause I was going to bed straight after school, so I didn't have time to do it. And then that lasted about half a term. And then at Christmas, something changed and everything sort of got a lot worse all at once. So the headaches got a bit worse and I stopped sleeping almost completely. So at nights where I was getting absolutely no sleep, there was nights where I was getting about two hours sleep. That was about the standard I was getting at that point. Uh, good night sleep was 4 hours then.’   ‘Participant: At the start it was headaches and fatigue and then the dizziness as well and difficulty thinking. Those all persisted, the sense of taste came like a day couple of days. UM. And then the dizziness got worse so there was a point where if I sat down and stood up, I'd have to wobble for a couple of moments before I'd be stable, there was a point where I stood up and I was like, OK, for a couple of seconds and then I just completely collapsed. But that might have been a medication that I was on at that point. The dizziness got better first. The headaches are still present. The fatigue is still present. The insomnia is still present. The brain fog is still present, but less, it's either not being triggered as much because I'm doing subjects that it likes more or it has, well, it has got a bit better too.’ |
| Abdominal pain or vomiting or nausea or reflux | - ‘Participant: uhh headaches, dizziness, nausea, umm sort of funny stomach gut sort of area that was never amazing umm but yeah the fatigue and the tiredness was the main one.’ - ‘Participant: I have recurring stomach pain.’   ‘Participant: because I just felt so sick I got an average of 2 or 3 hours a night.’  ‘Participant: because every time I shut my eyes it was just intense waves of nausea’  ‘Participant: so even my OT was saying she was struggling to keep up with all of my symptoms because a lot of them aren’t anything to do with chronic fatigue they’re purely to do with Long COVID so like when I had the gastro problems um I couldn’t eat anything or I ate but it was incredibly painful I was very weak I couldn’t lift my head off the sofa so there was no way I was going to be like practising climbing the stairs and going out for walks and building up my energy envelope when I couldn’t even stand up?’  ‘Participant: I’ve got acid reflux constantly recurring’   - ‘Participant: But I want to say around the 4th one is when the long covid really started to happen which was um not good um it was there was a lot of different symptoms other than just the normal ones from covid’ ‘Participant: stomach aches uh burning feet the reflux thing where my chest it was burning and it felt like it was going up and down my chest headaches um a lot of stuff like that.’ |
| Sensory issues | - ‘Participant: I had sensory difficulties with lights and smells and sounds.’   ‘Participant: I couldn’t do anything but watch a very specific type of youtube video because some noises were too loud some lights were too bright.’  ‘Participant: we couldn’t have the TV on at all because of the noise the sensitivity.’   - ‘Participant: yeah it really hurts my eyes seeing bright lights.’ - ‘Participant: Or it tastes horrible.’ ‘Yeah. Or when you could taste things, they did taste, certain things tasted horrible. Well, at first, everything tasted horrible.’ ‘Participant: Like sick.’ - ‘Participant: At the start it was headaches and fatigue and then the dizziness as well and difficulty thinking. Those all persisted, the sense of taste came like a day couple of days. UM. And then the dizziness got worse so there was a point where if I sat down and stood up, I'd have to wobble for a couple of moments before I'd be stable, there was a point where I stood up and I was like, OK, for a couple of seconds and then I just completely collapsed. But that might have been a medication that I was on at that point. The dizziness got better first. The headaches are still present. The fatigue is still present. The insomnia is still present. The brain fog is still present, but less, it's either not being triggered as much because I'm doing subjects that it likes more or it has, well, it has got a bit better too.’ |
| Brain fog | - ‘Participant: I’ve got and brain fog still’ - ‘Participant: At the start it was headaches and fatigue and then the dizziness as well and difficulty thinking. Those all persisted, the sense of taste came like a day couple of days. UM. And then the dizziness got worse so there was a point where if I sat down and stood up, I'd have to wobble for a couple of moments before I'd be stable, there was a point where I stood up and I was like, OK, for a couple of seconds and then I just completely collapsed. But that might have been a medication that I was on at that point. The dizziness got better first. The headaches are still present. The fatigue is still present. The insomnia is still present. The brain fog is still present, but less, it's either not being triggered as much because I'm doing subjects that it likes more or it has, well, it has got a bit better too.’ |
| Appetite loss | - ‘Participant: for the first couple of months all I ate all day was like a bowl of soup and some crackers.’ - ‘Participant: uh yeah, I wasn’t getting out as much as I used to because I was either too tired or just not feeling up to it and it affected my eating habits and I just kind of stopped doing a lot of the things that I used to do.’ - ‘Participant: it’s like I’m eating at really random times of the day, sometimes I can eat a lot in the day but not often and other days its like I just eat my tea at night and even at school like snacks and all of that but sometimes I can not eat any of it and then it’s like sometimes I have those ready meals don’t I and I can barely eat half of that.’ |
| Changes to menstruation | - ‘Participant: Yes so I was on, well ever since I started my periods which was year 7 September I had about 4 months which were regular like ish for starting a period and then during the march where lockdown started, the first lockdown, I was bleeding everyday for about 180 days. Participant: Umm yes so then that was the whole thing when we went to hospital because I was so unwell, was it the long COVID having a play on it or was it the other thing, you just didn’t know, now we think it was the long COVID because the pills, when we went to the gynaecologist, umm, they were saying the pills that I’m on wouldn’t cause fainting and dizziness so it’s got to be down to the long COVID’ - ‘Participant: On top of that my periods are bad so I have like a week where I’m basically incapacitated because of them and COVID’s changed my periods as well.’ |
| Mental health | - ‘Interviewer: Lovely, because you were saying that when you came out of hospital they were wondering if you were having some difficulties with your mental health, is that something you’d experienced before having COVID or was that a new thing?’ ‘Participant: well sometimes I could feel really down like that could happen, it still happens a bit now when I’m just really fed up and just want to lie in bed forever’ ‘Interviewer: yeah okay and with the Long COVID has that kind of got worse or has it stayed the same?’ ‘Participant: Well a lot more often I’m really upset about it and whenever I’m ill off school I would start crying because I just really want to get out back to normal but then it’s like I have this problem and everything yeah and a lot of the time’ ‘Participant: yeah’ ‘Participant: And we it has taken a toll on my mental health, and I don't really socialise an awful lot.’ |
| Blackouts | - ‘Participant: And there was less socialising, for because obviously I had COVID I got quite ill with the fact of blacking out quite a lot.’   ‘Participant: And there was less socialising, for because obviously I had COVID I got quite ill with the fact of blacking out quite a lot. |
| Vision problems | - ‘Participant: I have vision problems caused by covid nothing serious just I need my glasses changing and so I had glasses before COVID but I need them stronger now.’ |
| Vertigo | - ‘Participant: yeah I had vertigo from the 1st one’ |
| Fever | - ‘Participant: I was in shorts and a T shirt for about a year and a half because I couldn’t manage having anything long sleeved and my temperature was raised for pretty much a year so I had low grade fevers for about a year.’ |

### Supporting quotations for themes developed

Table 6: Supporting quotations for themes developed

| Theme name | Supporting quotations |
| --- | --- |
| Validation of experience. | **Subtheme 1) Young people appreciated healthcare professionals who had knowledge of Long COVID.**  **Interview F)**  ‘Participant: I don’t dislike anything I thought it was great.’ ‘Participant: Yeah.’  **Interview A)**  ‘Interviewer: um and how have you found the like people you’ve interacted with in the clinics kind of interacting with them how’s it been has it been okay?’ ‘Participant: Yeah it’s been fine easy to talk to um and very understanding of the situation which is the difference of speaking to someone whose specialised in it over the past couple of years to just a general doctor or GP.’  **Interview K)**  ‘Interviewer: And what else do you like about it?’ ‘Participant: Um I kind of like being explained some of the stuff and I like being able to talk to someone about it who knows.’ ‘Interviewer: Does it feel like they understand?’ ‘Participant: Yeah.’ ‘Interviewer: Which I can imagine is really helpful.’ ‘Participant: Yeah it is.’  **Interview B)**  ‘Interviewer: That's a long time. Very long time. And then when you had your first appointment in the ME CFS clinic, how did you feel after that?’ ‘Participant: Yeah, we did. I think it was. I think it was just because the doctors had been so kind of dismissive it’s all in your head. There's not a lot. Everyone goes through it. And I was suffering with pain every day and I still am. And it's like for a while I found that really hard and like even when we moved, I found it really hard just to be able to go to the doctors because I was just like, I don't want them to be telling me it's all in my head again. So, when we had the doctors or the CFS people turn around and go, oh my God, you've been through so much.’ ‘Participant: Yeah. And also, just saying like.’ ‘ Yeah. And they said they, you've been through so much and we get you've got through this and like you're X. I was X. Yeah, X at the time. And they. Like, I can't believe that you've still been able to kind of cope with this at the time you're in it. And so, it was like it. It made us, it made me cry at the end of it because I was very much like, I felt, listened to, and I felt like there was going to be at least someone that's going to kind of look into making a change.’ ‘Interviewer: Yeah, you felt heard. You felt understood. You felt like someone was doing something to help you, which is such a nice feeling.’  ‘Participant: Physical the fatigue and the mental health and just being able to talk about the mental health part has also helped, yeah, without feeling judged. It's like such a huge thing.’  **Interview I)**  But we definitely needed the clinic. We wouldn't have been able to get here without the support they gave with school, the management strategies and just the sort of connection to talk to someone who knows what's going on.’  **Subtheme 2) Young people were validated in the knowledge that other CYP were using the Long COVID clinic**  **Interview F)**  ‘Interviewer: And what was that like for you X? knowing that you weren't alone?’ ‘Participant: Um, you do. It does get you down obviously like, when you're not able to do things you are before it did. I was really upset. Um. And then, just to hear that I'm not the only one going through it like there are other people just don't worry. It was just like it was like a weight off the shoulder.’  **Interview C)**  ‘Interviewer: What do you think would be most helpful for you then, what kind of support are you looking for?’ ‘Participant: um I think um obviously hospitals should have areas, because we don’t really have a Long COVID area in the hospital do we?’ ‘Participant: Which I think that would be a better idea to be able to go there and actually face to face other people who have it and all go together and tell each other things like help and what not.’ ‘Interviewer: What like other young people?’ ‘Participant: Yeah.’ |
| Young people wanted to be proactive in their Long COVID management | **Interview E)**  ‘Interviewer: yeah fair enough and what have you liked about the Long COVID clinic?’  ‘Participant: umm I liked how they kind of given me lots of information because it’s nice knowing that you know all these things can be ruled out and these things might help it’s quite relieving to know that people are you know doing something.’  **Interview B)**  ‘Participant: And so, it was like it. It made us, it made me cry at the end of it because I was very much like, I felt, listened to, and I felt like there was going to be at least someone that's going to kind of look into making a change.’  ‘Interviewer: Yeah, you felt heard. You felt understood. You felt like someone was doing something to help you, which is such a nice feeling.’  **Subtheme 1) Regular clinic appointments helped young people manage their symptoms of Long COVID**  ‘Participant: sometimes I can still feel very exhausted, but it’s definitely helped and it has calmed me down a lot because sometimes I get very stressed wondering how long this is going to go on for.’ ‘Interviewer: yeah that must be really tricky.’ ‘Participant: but it’s the fact that every two weeks I’m up here it calms me down a lot.’  **Interview B)**  Participant: And also, it's quite nice just to like when you kind of go because we've got a shorter period of meetings now, so instead of doing it every two weeks, it's every like every one week instead and like working with that, and just kind of being able to, if I had a question that week and trying to want to change something or maybe improve something else. Well, given like understandings to try and make myself understand it more. It's a lot more helpful and when I'm struggling with something that's going on in my mind and it's feels like it's a kind of circle with the fact that I have the fatigue and then I have the mental health and then I have the physical health and it's just like it's just and then the taste and everything going around in a circle, you kind of like get trapped and sometimes it's nice to yeah, have that support; when like if it was the week that I wasn't talking to her, it would be quite hard to her and go, oh, this happened last week, but being able to have it and go, this is happening right now. It helps me kind of move forward a bit more.’  **Interview H)**  ‘Interviewer: Okay that’s good. And you were given that energy management plan, how regularly would you say you were like having meetings?’ ‘Participant: Every couple of weeks.’  ‘Interviewer: Oh that’s good. That’s good. And was it always like with the same person?’ ‘Participant: Yeah’ ‘Interviewer: Yeah, nice. And did you find regular check in contact helpful, would you like to have like more less?’ ‘Participant: I thought it was really good, the amount I had. And just I feel like I even now if I need to have another meeting I can, I feel like I’ve got kind of that support there.’  **Subtheme 3) Learning about Long COVID helped young people manage their symptoms of Long COVID**  ‘Interviewer: yeah definitely definitely so much better to have those specialist input kind of thing um so would you recommend the clinics overall with what you’ve had so far to other people?’ ‘Participant: yeah I think definitely if your someone who wants to be open about it and talk about it and even if it’s just to discover more for yourself what the whole thing is I think it’s a good thing to do’  **Interview E)**  ‘Interviewer: yeah fair enough and what have you liked about the Long COVID clinic?’ ‘Participant: umm I liked how they kind of given me lots of information because it’s nice knowing that you know all these things can be ruled out and these things might help it’s quite relieving to know that people are you know doing something’ ‘Interviewer: yeah definitively and although you’ve seen lots of different types of people which can be quite confusing sometimes have you liked that you’ve met different types of people like psychologists, physiotherapists, people like that’ ‘Participant: yeah I think I have quite a bit because it’s nice having lots of people with different **skillsets** so it’s nice you know having a wide range of information’  ‘Interviewer: yeah, and X how have you felt about all these appointments with different type of people?’ ‘Participant: well I mean I found it quite interesting to hear what they all you know what they think and what they can see and what it might be but I’ll be honest after the especially long ones like hour hour and a half ones it is a little bit I feel a little bit tired afterwards but I think I mean its worth feeling that tired finding out the information.’  **Interview K)**  ‘Interviewer: And what else do you like about it?’ ‘Participant: Um I kind of like being explained some of the stuff and I like being able to talk to someone about it who knows.’  ‘Interviewer: It’s sounds like it’s been quite helpful.’ ‘Participant: Yeah they were helpful and they also did explain quite a lot to me.’  **Interview B)**  ‘Interviewer: What else has been helpful about being a part of the MECFS clinic? Is there anything else that you can think of?’  ‘Participant: Yeah. So, a lot of the time they kind of send like emails and debriefs after it because I obviously suffer from brain fog and so I have mum in the lot of them, because like she remembers it all and also I have a bit of anxiety on things like this. So, she kind of helps me kind of relax to the point of able to talk. Yeah so it's not been too bad, but like they've kind of helped kind of digest it after and give the leaflets to kind of go through PowerPoints that explains certain bits for like my age or what we could make better so yeah, that's definitely been a big help.’ |
| Young people desired normality | **Subtheme 1) Young people valued flexible appointments to help have a sense of normality**  **Interview A)**  ‘Interviewer: that’s good did they kind of like offer you times or did you tell them when was best for you?’ ‘Participant: um it was a bit of both so they sort of like said what they like prefer or what they think would be realistic and then if it wasn’t going to work we’d reply and say like this is how we can fit it in with how we’re feeling or like what we’re doing it was quite lenient quite easy I’d say to fit in an appointment once we had one booked in.’  **Interview G)**  ‘Interviewer: did you think the appointment times worked with your schedule because I know you said you had other clinics things like that? Participant: yes yeah they did and we planned it and they’ve always kept it in the afternoon which is really helpful for me.’  **Interview H)**  ‘Interviewer: would the appointment times did they like work with your schedule, did they kind of like cater it around you or did you have to like miss school anytimes?’ Participant: Yeah it was all catered around me, so whenever I had free periods I would organise one. Basically we just planned the date and time that suited us both and it always worked. I didn’t miss any school for it.’  **Subtheme 2) Young people valued online appointments to help with their Long COVID management**  **Interview G)**  ‘Interviewer: that’s good that they were flexible like that.’ ‘Participant: um yeah online I can’t get there so far.’ ‘Interviewer: did you think that it was better being online then?’ ‘Participant: yes’.  **Interview L)**  ‘Interviewer: and how easy was it for you to get to your clinic appointments, because obviously they were on zoom, did you find it easier that they were online, or would you have preferred some in person sessions would you say?’ ‘Participant: online was easier because I did have school and having to go to a physical place would have just made it a lot harder so zoom was fine.’  **Interview H)**  ‘Interviewer: That’s good. And what did you think about it being online?’ ‘Participant: Um yeah it was fine, I feel like I’m used to that, I was used to online stuff by then.’ Interviewer: Would you in a way rather it have been in person or?’ ‘Participant: Not I’m yeah, I don’t think I would have gained any, I think it would have been worse because I would have had to travel for it and then it would have been a bigger thing and probably more energy consuming.’  **Interview I)**  ‘Participant: UM. The online appointments are really helpful. If I'd had to travel to X because I'm up in the X, yeah, that would have triggered my fatigue before I even got to the appointment. So don't stop the online appointments because it just makes it so much easier if you're on one end of the country, you don't have to drive all the way to the other end to get advice.’ ‘Interviewer: Is there anything you don't like about it being online or is it just good overall?’ ‘Participant: Not particularly.’  ‘Interviewer: Yeah. You still feel like you get the same out of the sessions?’ ‘Participant: Yes. Because it's not like you're doing a physical exam. So, you don't need to really be there in person, but with fatigue it's much easier if you're in an environment you're comfortable with, with people that you're comfortable with, that's close. So, you don't have to travel hundreds of miles. And it just makes the whole process much easier.’  **Subtheme 3) The clinics helped the young people gain reasonable adjustments for school**  **Interview E)**  ‘Interviewer: That’s good do you think as time has gone on and we’re talking a bit about Long COVID more it’s become easier to talk about with school and with other people?’  ‘Participant: It is yeah because I mean from the appointments my mum gets some letters sometimes and she forwards them to the school which means they have all the information and it means if there’s any like really big issues I know who to talk to yeah.’  ‘Interviewer: Yeah that’s really good.’    **Interview I)**  ‘Interviewer: So, like where you are now, would you say that overall, the Long COVID management plans have improved your symptoms, or would you say it's kind of got better with time? How are you now kind of thing?’  ‘Participant: I'd say a bit of both. The CBT was really helpful with getting through my exams. I wouldn't have been able to do those without the management strategies that that gave me. But that I don't think helped me get better overall, that just helped me manage what I had at that point and continue through that to do what I had to do. I have got better over time, partly because we've done research and found things that have worked and partly just because time. But we definitely needed the clinic. We wouldn't have been able to get here without the support they gave with school, the management strategies and just the sort of connection to talk to someone who knows what's going on.’ |
| Waiting for versus getting help | **Subtheme 1) The wait to access Long COVID clinics was too long**  **Interview A)**  ‘Interviewer: is there anything else you like about them you said it’s good to be able to talk about things and is there anything you find is particularly helping?’  ‘Participant: um I mean I probably would more if it happened like 6 months to a year ago when I was properly run down with it but now that I’m coming out the end of the Long COVID um I don’t think there’s as much sort of help needed if that makes sense’  ‘Interviewer: um hum yeah’  ‘Participant: So it doesn’t feel like its benefiting as much as it could.’  ‘Participant: Umm well to be honest we’ve only really started seeing specialists with Long COVID or fatigue clinics in the last I’d say few months really which is typical really just as I’m starting to feel better um because which I mean fair enough to them there was like not enough knowledge about COVID it’s all very new and like there’s not much that could really have been done.’  ‘Interviewer: Yeah.’  ‘Participant: It was just to have that like access to support like in general would have been nice because there really wasn’t much offered over the past two years and it’s only really recently that’s stuffs been offered out more and there’s still big queues to get sort of any help and advice.’  ‘Interviewer: Yeah when did you kind of first seek help for the Long COVID then?’  ‘Participant: Umm we were seeking it pretty much like late 2020 I guess autumn winter of 2020 was when we started looking for help or like trying to figure out more of it and then going into 2021 we were looking for help and then yeah it was just this year really last few months that we managed to receive proper help for it.’  ‘Interviewer: yeah okay and then so how long were you kind of left without support I know it would be very difficult to say specifically but from the onset of the long covid till when you started accessing proper clinics how long roughly do you think that window was?’  ‘Participant: Um for proper like chronic fatigue clinic it would be roughly two years I’d say’  ‘Interviewer: Okay yeah so kind of um would you say that overall the clinics you have found them quite helpful but they’ve come in quite late in your journey?’  ‘Participant: Yeah exactly that.  ‘Interviewer: And now you’re nearly all better kind of thing.  ‘Participant: So if I’d got the help I received in the last couple months about a year ago it would have been like great like perfect but now it’s just a little bit too long.’  ‘Interviewer: Yeah did the doctors the GPs know about the clinics just out of interest?’  ‘Participant: Um I think so but I think they were hesitant to refer us to them because A just the queues were massive so even once we did get referred we were waiting in the queues for quite a long time to get seen and then also there were just like conscious not to override them seeing how many people were being sent there anyway so they tried to help as much as they could without knowing much about it.’  **Interview G)**  ‘Participant: Kind of I’m relatively new to the hub because that took ages to get through aswell and then I needed a plethora of blood tests um and then they didn’t see us for ages because obviously there’s a lot of children with Long COVID now.’  ‘Participant: Obviously the referral to the chronic fatigue service or the long covid clinic as you know it if I call it the chronic fatigue service I’m sorry it’s because I’ve got so many different clinics that I’m under the chronic fatigue service had to be done through the GP and my GP is not known for being the best GP in the area and they take about they tried to refer me to the adult service twice and then they couldn’t find the information for the kids service so they just gave up and didn’t tell us for 4 months’  ‘Interviewer: Gosh.’  ‘Participant: So we didn’t have any help for 4 months and then we had to go back to the consultant who then spoke to the manager I think and sort of said right we need to get this girl in so he ended up doing it instead.’  ‘Participant: There’s a lot of children with covid and there’s a lot more children going down with Long COVID you could be safe in your 1st 2nd 3rd infection and then get covid on the 4^th^’ ‘Interviewer: It’s so random yeah.’  ‘Participant: And suddenly need the service so when I was there the queue was a lot shorter so it did take 4-5 months only because my GP didn’t have my information in the system’ ‘Interviewer: yeah yeah more of any issue with the GP kind of side of things’ ‘Participant: It was more of an issue of the GP than it was of the clinics.’  **Interview J)**  ‘Interviewer: The whole journey that you have been on, what would make that better for you?’  ‘Participant: If they could like figure out how to make it go a bit faster, because I had to wait years before even getting referred.’  **Interview K)**  ‘Interviewer: So tell me how easy it was to get an appointment to see a health professional at the Long COVID clinic?’  ‘Participant: Um I think it was a year.’  ‘Interviewer: Yeah that is quite a long time.’  ‘Participant: Yeah.’  ‘Interviewer: How did you feel about waiting all that time to get that appointment?’  ‘Participant: Well it was a bit frustrating.’  ‘Interviewer: Can you think of anything that could make the Long COVID clinic better?’  ‘Participant: Um not really maybe um maybe not such a long waiting list.’  **Interview M)**  ‘Interviewer: OK. And (participant), what is that process been like for you, like waiting to try and actually see someone about how you're feeling?’  ‘Participant: Long.’  ‘Interviewer: Is there anything that you have disliked about the service so far.’  ‘Participant: It's quite long to be fair.’  ‘Interviewer: Yeah. And when you say it's quite long, is it because you've had to wait so long to get to see the service or is it because the whole process is really long and you want a quick fix process, yeah.’  ‘Participant: Process. That's it. And I think the service, how long have we been waiting?’  **Interview H)**  ‘Interviewer: How did you discover the Long COVID services?’  ‘Participant: I think my mum looked into it we went to the my GP uh just to talk about it just having Long COVID and my mum was like “I actually read about this thing” and he was like “yeah I can sign you up” so we did and then it took a while to get in because I don’t know it took a while to get in because something messed up but then I got in and yeah.’  ‘Interviewer: Oh, okay and how long would you say roughly did it take from going to the GP to starting your first appointment?’  ‘Participant: I remember I think I started at the beginning of year 11, and a couple of months, I remember it being like a while because I remember my mum being annoyed.’  **Interview I)**  ‘Participant: Because it's always really busy, so it's like six months to get an appointment with the pediatrician, and then we only see him once every six months, and every time we have to convince him it's actually Long COVID again. And then it's another 3-6 months to get referred to the fatigue clinic and then that's another couple of months to get onto the CBT. So, it doesn't feel like there's enough people doing it for the amount of people who need it, which is no fault of the doctors.’  **Subtheme 2) Young people wanted information on how to help manage their symptoms whilst waiting for an appointment**  **Interview J)**  ‘Participant: um I’ve only had one meeting with the clinic so far and I have to wait like another couple of months and I’m on another waiting list to see them’  …..  ‘Interviewer: So have they given you some things to do while you wait for your next appointment? ‘ ‘Participant: No’  ‘Interviewer: But they have said to you it would be something to do with energy management and breathing techniques?’  ‘Participant: yeah’  ‘Interviewer: and how do you feel about that do you know much about these things?’  ‘Participant: um well I have asthma so I know quite a bit about the breathing exercises but yeah I don’t really know about anything else.’  **Interview I)**  ‘Participant: UM. Well, before we sort of got round to the fatigue clinic. For treatment with fatigue, there wasn't sort of at least that I know of any like NHS page with recommendations. There might be, and I just haven't found it, but we didn't really know what to do with fatigue management. We didn't know what to do with CBT. We didn't know how to manage my dizziness or headaches or any of my symptoms, really.’  Participant: Yeah. And also, just general tips and advice, so don't overdo it. Don't get into a boom-and-bust cycle. And when you're doing too much and then you crash and repeat over and over again. Like ways to help yourself rest like meditation or CBT or anything. As far as I'm aware, I might be incorrect in this, but there is no place that you can go where all that information is sort of in one place on the NHS website. But I might be incorrect on that.’  **Subtheme 3) Young people wanted the clinics to communicate with their school whilst they were waiting**  **Interview I)**  ‘Participant: Public or if there was someone you could contact to get those things, because if you like, got Long COVID couple of months before your exam, the chances that you're going to get seen, see if specialist in that time before your exam to be able to take that exam with the mitigating circumstances are very slim, so either you don't sit your exam, or you sit it and really really struggle.  Interviewer: Yeah, there needs to be like a way to get the support for school quicker.’ |
